# Gastric coagulation and postprandial amino acid absorption of milk is affected by mineral composition: a randomized crossover trial

**DOI:** 10.1101/2023.09.13.23295475

**Authors:** Elise J.M. van Eijnatten, Julia J.M. Roelofs, Guido Camps, Thom Huppertz, Tim T. Lambers, Paul A.M. Smeets

## Abstract

**Background:** *In vitro* studies suggest that casein coagulation of milk is influenced by its mineral composition, and may therefore affect the dynamics of protein digestion, gastric emptying and appearance of amino acids (AA) in the blood, but this remains to be confirmed *in vivo*.

**Objective:** This study aimed to compare gastrointestinal digestion between two milks with the same total calcium content but different casein mineralization (CM).

**Design:** Fifteen males (age 30.9±13.8 y, BMI 22.5±2.2 kg/m2) participated in this randomized cross-over study with two treatments. Participants underwent gastric magnetic resonance imaging (MRI) scans at baseline and every 10 min up to 90 min after consumption of 600 ml milk with low or high CM. Blood samples were taken at baseline and up to 5 hours postprandially. Primary outcomes were postprandial plasma AA concentrations and gastric emptying rate. Secondary outcomes were postprandial glucose and insulin levels, gastric coagulation as estimated by image texture metrics, and appetite ratings.

**Results:** Gastric content volume over time was similar for both treatments. However, gastric content image analysis suggested that the liquid fraction emptied quicker in the high CM milk, while the coagulum emptied slower. Relative to high CM, low CM showed earlier appearance of AAs that are more dominant in casein, such as proline (MD 4.18 µmol/L, 95%CI [2.38-5.98], p<0.001), while there was no difference in appearance of AAs that are more dominant in whey protein, such as leucine. The image texture metrics homogeneity and busyness differed significantly between treatments (MD 0.007, 95%CI [0.001, 0.012], p=0.022; MD 0.005, 95%CI [0.001, 0.010], p=0.012) likely because of a reduced coagulation in the low CM milk.

**Conclusions:** Mineral composition of milk can influence postprandial serum AA kinetics, likely due to differences in coagulation dynamics.

## INTRODUCTION

Protein is an essential macronutrient for many functions in the human body^1, 2^. Consuming sufficient protein can be a challenge. Therefore, optimal digestion and absorption of the consumed protein support bioavailability of amino acids (AA)^3, 4^. A common source of protein is bovine milk^5^, which generally contains about 3.5% protein, of which caseins represent around 80% and whey proteins around 20%^6^. While whey protein remains soluble, casein coagulates in the stomach when casein micelles are destabilized by pepsin proteolysis^7^. This leads to the formation of curds containing protein and fat globules that impact overall digestion kinetics, such as protein digestion and absorption^8^.

Previous studies, predominantly *in vitro*, suggest that casein coagulation is affected by several factors, including processing-induced protein modifications, overall product composition, including mineral composition, and variations in gastric acidification and protease secretion^9–12^. It is important to fully understand the effect of processing-induced protein modifications of milk on coagulation since coagulation could influence the rate at which protein empties from the stomach and thereby affect protein digestion and absorption kinetics^13^. Gastric emptying (GE) is the rate-limiting step in the delivery of nutrients to the small intestine for further breakdown and absorption^14^. GE rate of liquids is mainly driven by energy density and viscosity^15^. However, compared to whey proteins, the coagulated casein fraction of milk empties later, because the stomach only empties particles sized below 1–2 mm^14, 16^. Accordingly, whey proteins, which remain soluble, have a higher GE rate than caseins^17, 18^. Processing of milk alters its functional properties including casein and its coagulation^19^, which could influence GE and related digestion kinetics. This is supported by recent *in vivo* work showing that the processing of casein and the resulting alterations in the product matrix can have a strong effect on postprandial AA responses. For instance, cross-linked sodium caseinate was more rapidly digested than micellar casein and calcium caseinate and upon the ingestion of dairy products containing 25 g protein, and a higher increase in EAA concentrations in blood was observed after consumption of yoghurt, compared to milk and cheese^20, 21^. The rate at which AA are absorbed greatly affects their availability for muscle protein synthesis^22^. A faster release of milk proteins results in a more prominent muscle synthetic response, whereas a slower release of protein from the stomach results in a better net postprandial protein utilization^7^. Thus, the degree of casein coagulation in the stomach could affect the dynamics of gastric protein digestion, stomach emptying, and subsequent intestinal digestion and absorption of AA.

Both the physical (e.g., compactness, hardness and elasticity, size of fat globules) and the chemical parameters (e.g., protein/lipid ratio, P/Ca ratio) can influence the milk matrix and could therefore affect the bioavailability of AA^3^. *In vitro* studies indicate that casein coagulation is affected by mineral composition^23^ since caseins form casein micelles with calcium, phosphate, and magnesium^24^. Partial decalcification of casein micelles results in looser-formed gastric clots and greater proteolysis^25^. Casein mineralization (CM) also affected the coagulation of milk proteins in a model infant formula^26^. The effect of CM on coagulation was followed up by a study on the coagulating behavior of bovine casein micelles under infant, adult, and elderly conditions where gastric coagula became looser and the formation of free amino groups and small peptides increased with an increasing level of decalcification^27^. However, thus far, only *in vitro* or indirect (by measuring AA kinetics) *in vivo* studies have been done, in which other product differences than only mineralization were studied. MRI could be a helpful tool in assessing casein coagulation. Currently, the main use of MRI in gastric research is measuring GE^28, 29^. But MRI can also be used to visualize intragastric processes, such as changes from liquid to solid state, gastric sieving and phase separation^30, 31^, which is an advantage over ultrasound or tracer-methodology. Since gastric protein coagulation involves a change from a liquid to a semi-solid state, MRI could potentially be used to quantify the degree of coagulation. So far, gastric coagulation has only been visually assessed using MRI^32^, however image texture analysis may provide a more objective and accurate quantification^29^.

This study aimed to compare gastrointestinal digestion (coagulation, GE, and postprandial AA dynamics) between skimmed bovine milks with the same total calcium content but a different degree of CM. We hypothesized that gastric protein coagulation would be more prominent in milk with higher CM and consequently delay gastric protein emptying, serum AA appearance, and related glycemic responses.

## PARTICIPANTS AND METHODS

### Design

This study was a randomized, single-blinded, crossover study with two treatments. Washout was at least one week, and sessions were a maximum two months apart. Primary outcomes were postprandial plasma amino-acid concentrations and gastric emptying rate. Secondary outcomes were postprandial glucose and insulin levels, gastric coagulation, and other product instabilities if visible and appetite ratings obtained after each MRI measurement. The results are presented in the order of (I) coagulation, (II) gastric emptying, (III) AAs, (IV) insulin / glucose and (V) appetite ratings.

### Participants

Healthy males, aged 18-55 years, BMI 18.5-25.0 kg/m^2^, were recruited from the Wageningen area from December 2020 to March 2021. Exclusion criteria were bovine milk allergy, lactose intolerance (either self-reported or diagnosed), gastric disorders or regular gastric complaints, such as heartburn, use of proton pump inhibitors or other medication which alters the normal functioning of the stomach, recreational use of drugs, within one month prior to the pre-study screenings day, alcohol consumption of more than 14 standard units/week, being vegan, smoking, or having a contraindication to MRI. Because there are sex differences in gastrointestinal function^33, 34^, males were chosen as the study group. The sample size was calculated for the first primary outcome, the postprandial AA profile, based on the expected difference in peak serum AA concentration. We estimated the peak difference to be 0.4 pmol/ml, with an SD of 0.3 pmol/ml based on an earlier study performed with differently treated dairy products that are comparable to these study products^21^. The power was set to 0.9. This resulted in an estimated sample size of n=14. However, to account for a possible smaller difference, it was decided to include 15 participants. Fifteen males (age: 30.9±13.8 years; BMI: 22.5±2.2 kg/m^2^) completed this study. See the flow diagram in **Supplementary figure 1**. The study was conducted according to the principles of the Declaration of Helsinki (October 2013) and registered with the Dutch Trial Registry under number NL8959 accessible through https://trialsearch.who.int/Trial2.aspx?TrialID=NL8959. All participants provided written informed consent.

### Study procedures

On the day before each test session, participants consumed a standardized 500-kcal rice dinner, which they could finish or eat less if they preferred. Drinks were free of choice. After this, participants fasted for at least 12 hours until the test session the following morning. Drinking water was allowed up to 1.5 hours before the session. Participants arrived at the hospital Gelderse Vallei, Ede, The Netherlands, at 8 or 10 AM and were measured at the same time on both study days. **Figure 1** shows an overview of a test session. Upon arrival, an intravenous (IV) cannula was placed in an antecubital vein. Baseline measurements consisted of appetite ratings, an abdominal MRI scan, and a blood sample. Subsequently, participants were instructed to consume 600 ml of a test product within five minutes, but all finished between one and four minutes. Gastric MRI scans were performed at baseline and every ten minutes, starting at T = 10 min up to 90 minutes after the start of consumption. During the MRI session, participants verbally rated subjective appetite: hunger, thirst, fullness, desire to eat, and prospective consumption on a scale from 0 (not at all) to 100 (most imaginable) at each time point^35^. Blood samples were drawn at baseline and at T = 15, 30, 45, 60, 75, 90, 120, 150, 180, 210, 240, 270, and 300 min.

**Figure 1.**
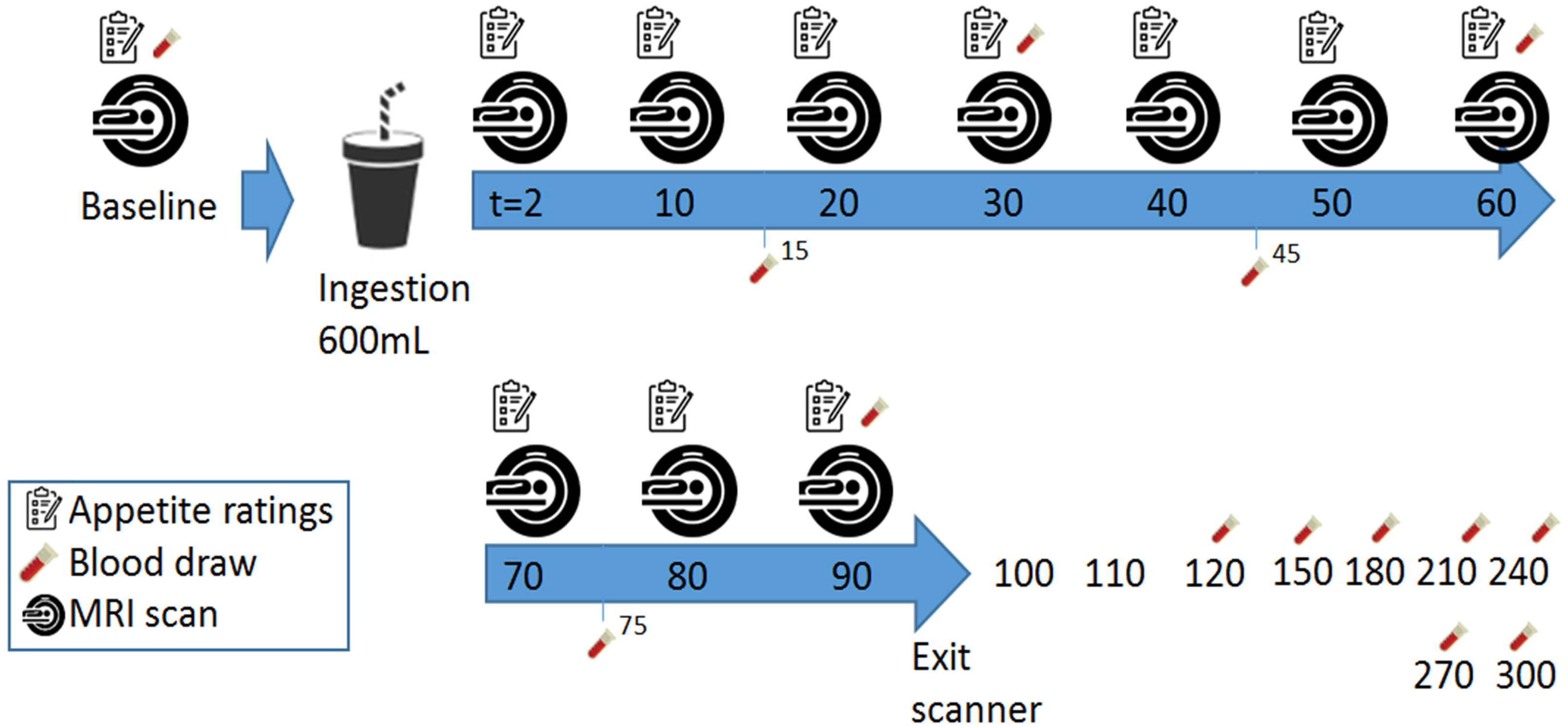
Overview of a test session

The test products were two skimmed milks with different micellar mineral composition, provided by FrieslandCampina. The low CM product was skimmed milk with 20 mM added trisodium citrate, resulting in a degree of CM of 4.3 mmol protein-associated Ca/10 g casein, determined as described previously^36^. Adding citrate alters the micellar calcium content and thereby the casein micelle integrity^37^. To maintain a similar buffering capacity in the high CM product, disodium hydrogen phosphate was added to skimmed milk at a level of 20 mM, resulting in a degree of CM of 8.9 mmol protein-associated Ca/10 g casein. Solutions were made at the same time in the afternoon preceding the test day and were slowly mixed with a magnetic stirrer at 4° C overnight to create an equilibrium of micellar and non-micellar calcium. The difference in gastric protein coagulation was determined by wet weight measurements of the coagulates during *in vitro* gastric digestion (**Supplementary figure 2**). The nutritional value of both test products per 600 ml prepared product was 831 kJ/195 kcal, 0 g fat, 27 g carbohydrate, 21 g protein and 762 mg calcium. Participants were randomly allocated by block randomization using randomizer.org to receive either the low CM or the high CM milk first. The milks were similar in appearance and taste and participants were blinded to the milk they received.

### MRI

Participants were scanned in a supine position with the use of a Philips Ingenia Elition X 3.0T MRI scanner. A T_2_-weighted ∼20-s 2-D Turbo Spin Echo sequence (37 4-mm slices, 2 mm gap, 1 x 1 mm in-plane resolution, TR = 550 ms, TE = 80 ms, flip angle: 90 degrees) was used with breath-hold command on expiration to fixate the position of the diaphragm and the stomach. The software Medical Imaging Processing And Visualization (MIPAV, version 11.0.3) was used to manually delineate gastric content on every slice^38^. Volumes on each time point were calculated by multiplying the surface area of gastric content per slice with slice thickness, including gap distance, and summed over the total number of slices showing gastric content. To assess changes in gastric coagulation, image texture analysis of the stomach content was performed using the software LIFEx (version 7.2.0, Institut national de la santé et de la recherche médicale, France)^39^. Homogeneity, coarseness, contrast, and busyness were calculated. These image metrics provide information on the spatial patterns of voxel intensity^40^. The Gray-Level Co-occurrence Matrix (GLCM) method was used for homogeneity (degree of similarity between voxels) and Neighborhood Gray-level Difference Matrix (NGLDM) difference of grey-levels between one voxel and its 26 neighbors in 8 dimensions was used for contrast (local variations), coarseness (spatial rate of change in intensity) and busyness (spatial frequency of changes in intensity). The number of grey levels for texture metric calculation was set at 64, intensity rescaling at relative (ROI: min/max), and dimension processing at 2D. On each postprandial time point, texture metrics were calculated per slice for the stomach content. Subsequently, a weighted average texture metric was calculated based on the gastric content volume in each slice such that slices with little stomach content contributed less to the average than those with more stomach content. To quantify the (relative) volume of liquid and semi-solid stomach contents the number of lighter (more liquid), intermediate and darker (semi-solid) voxels was calculated by determining intensity thresholds with the use of Otsu’s method^41^ in Matlab (version R2023a, multitresh function) an approach previously used on *in vitro* MRI images of milk digestion^42^. The number of intermediate and darker voxels were summed and interpreted as reflecting coagulation. This was done because visual inspection of the thresholding results showed that in these images a separation in two categories was not accurate. In the context of this study, changes in image texture metrics were interpreted as reflecting changes in the degree of coagulation. An example of stomachs with and without coagulation and their corresponding image texture measures can be found in **Supplementary figure 3**.

### Blood collection and analysis

Blood samples (10 ml) were drawn from the IV cannula into sodium-fluoride, serum-, and lithium-heparin tubes. After collection, sodium-fluoride and lithium-heparin tubes were centrifuged at 1000 x *g* for 10 min at 22°C within 30 min, to obtain blood plasma. To obtain blood serum, serum tubes were first allowed to clot for 30 min before being centrifuged at the same conditions as the other tubes. Following centrifugation, serum aliquots of 500 µl and 250 µl were pipetted in 5 ml tubes and stored at -80°C until they were analyzed in bulk.

Analysis and quantification of serum AA concentrations were done using liquid chromatography mass spectrometry (LC-MS) triple quad, with an internal standard and 13C reference mixture^43^. For determination of the glucose concentrations, plasma samples were processed using an Atellica CH Glucose Hexokinase_3 (GluH_3) assay kit and quantified using an Atellica CH analyzer (Siemens Healthineers, The Netherlands). The lower limit of detection was 0.2 mmol/l and intra-assay CVs were at most 4.5%. Serum insulin was processed and its concentrations were quantified using an enzymatic immunoassay kit (ELISA, Mercodia AB, Sweden) with a limit of detection of 0.008 mmol/l and intra assay CVs of at most 6.8%.

### Statistical analysis

To estimate gastric emptying half time (GE-t50), a commonly used summary measure for each scan session, a curve was fitted according to an established linear-exponential model as developed based on earlier models of GE to the data of gastric volume over time using R statistical software^16, 44–46^. This method works well for gastric content that increases due to gastric excretion in the early phase (lag phase) and afterward empties almost linearly. Further analyses were performed in SPSS (version 22, IBM, Armonk, USA). GE-t50 was compared between low and high CM milk with a paired t-test. The serum AA were categorized into three groups: branched-chain amino acids (BCAA), essential amino acids (EAA), and non-essential amino acids (NEAA) and their content was summed. Overall blood parameters, gastric volume, image texture metrics, and appetite ratings were tested using linear mixed models with treatment, time, and treatment*time as fixed factors and baseline values as covariate. An extra analysis on the first 90 min of the blood parameters was conducted since this is when differences in gastric coagulation were expected. Proline, an AA more dominant in casein, and leucine, an AA more dominant in a whey protein, were used as a showcase since the expected difference in AA kinetics would be driven by differing casein coagulation and not whey. After this, we estimated the contribution of casein and whey proteins by determining the casein/whey AA ratio ‘Q’ of serum AA from the concentrations of proline, phenylalanine, aspartic acid, asparagine, and alanine according to the method of Jacobs et al. ^47^ described for food products. Normality was confirmed by quantile-quantile plots of the residuals, except for insulin, which was log-transformed to meet normality. Missing data was handled using a Maximum Likelihood estimation. The significance threshold was set at p = 0.05. Data are expressed as mean ± SD unless stated otherwise.

## RESULTS

### Coagulation metrics

Qualitatively, coagulation was visible on the MRI images for both high and low CM milk (**Supplementary figure 4**). The image texture metrics homogeneity and busyness were higher for high CM milk (MD 0.007, 95% CI [0.001, 0.012], p=0.022 and MD 0.005, 95% CI [0.001, 0.010], p=0.012, respectively). Coarseness (MD 0.001, 95% CI [-0.001, 0.002], p=0.512),) and contrast (MD -0.086, 95% CI [-0.203, 0.031], p=0.149) were not significantly different. There were no time*treatment interaction effects. **Figure 2** shows homogeneity over time as an example. Figures of busyness, coarseness, and contrast can be found in **Supplementary figure 5.**

**Figure 2.**
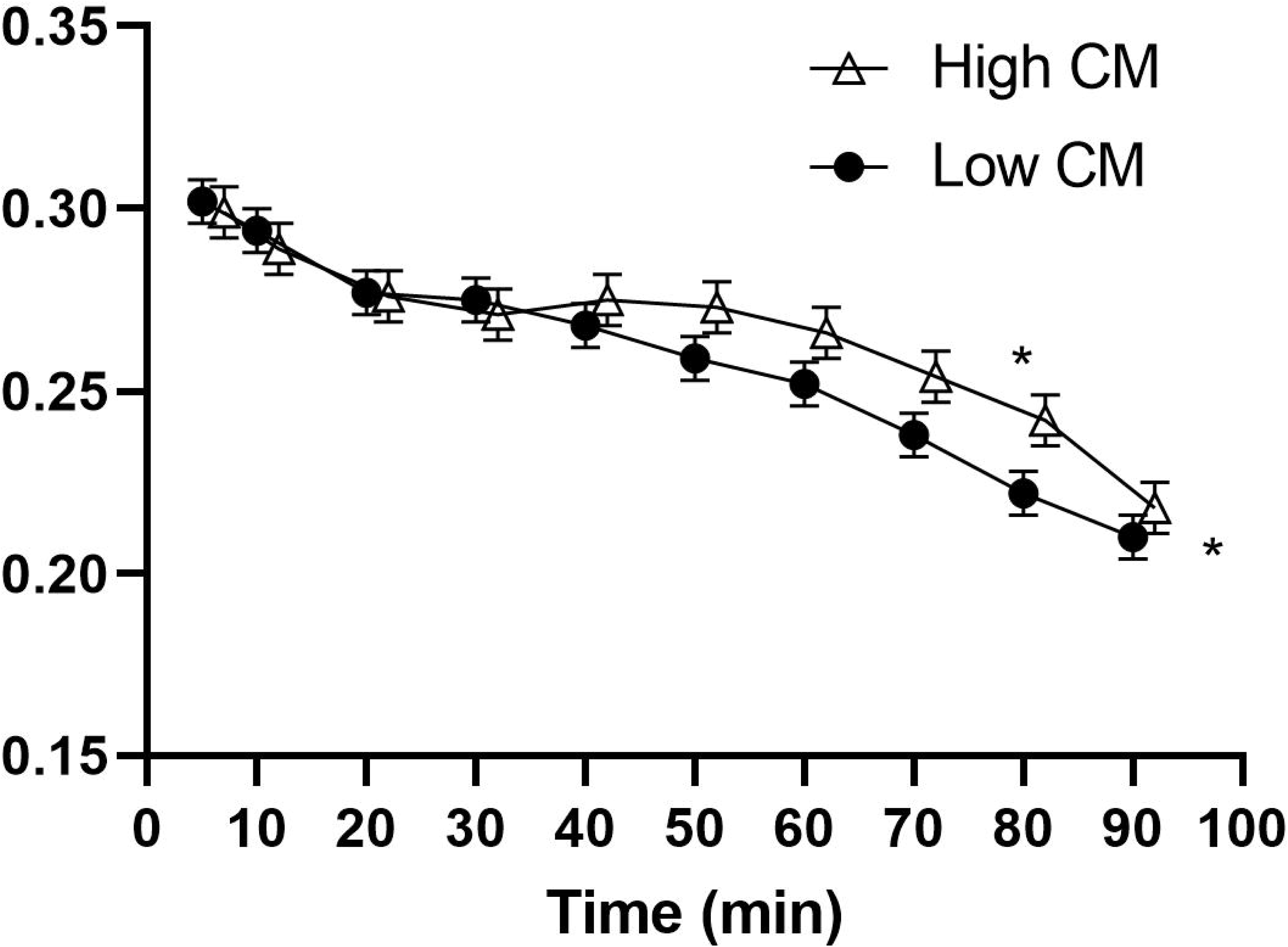
Mean ± SEM of image texture metric homogeneity of gastric content as visible on MRI after low and high CM milk ingestion. *p < 0.05 placed above the value denotes a significant time point, at the right of the graph it denotes a significant treatment effect. A difference in image texture metrics may reflect a difference in casein coagulation.

### Gastric emptying

There was no significant difference in gastric volume over time between treatments (MD 3.8, 95% CI -8.2, 15.8, p = 0.53). This is in line with the GE-t50: low CM milk 45.6 ± 7.8 min and high CM milk 46.6 ± 8.7 min (MD 1.0, 95% CI [-1.9, 4.0], p = 0.46) (**Figure 3**). There was a higher proportion of liquid over time between treatments in the low CM condition (MD 2.2 %, 95% CI [0.30, 4.1], p = 0.023) (**Supplementary figure 6**).

**Figure 3.**
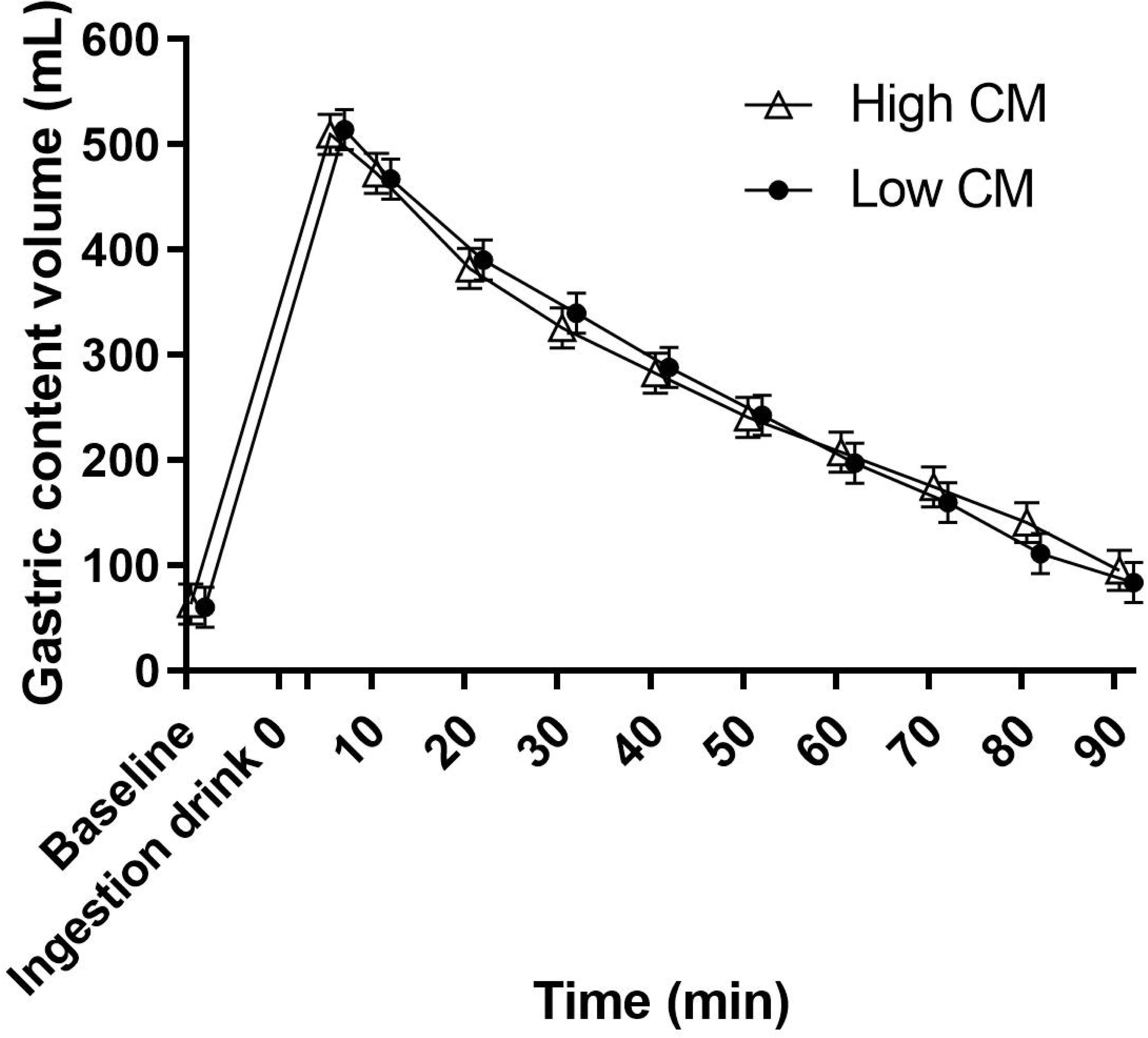
Mean ± SEM gastric content over time after ingestion of 600 ml of low and high CM milk. There were no significant differences between the two treatments.

### Amino acids

The total EAA postprandial response over time was similar for low and high CM milk (MD -2.6 µmol/L, 95% CI [-8.5, 3.2], p = 0.379, time p < 0.001, time * treatment p = 0.374) and total NEAA response over time was higher for the low CM milk with a trend for the interaction of time and treatment (MD µmol/L -4.3, 95% CI [-7.4, -1.1], p = 0.008, time*treatment p = 0.095), driven by time point T = 60 min, p<0.001 (**Figure 4**). The BCAA valine (MTD -1.784, 95% CI [-3.531, -0.038], p = 0.045, time p < 0.001, time* treatment p = 0.135) driven by t = 60 min, p = 0.001) was significantly higher for the low CM milk. Figures of separate AAs can be found in **Supplementary figures 7, 8 and 9**.

**Figure 4.**
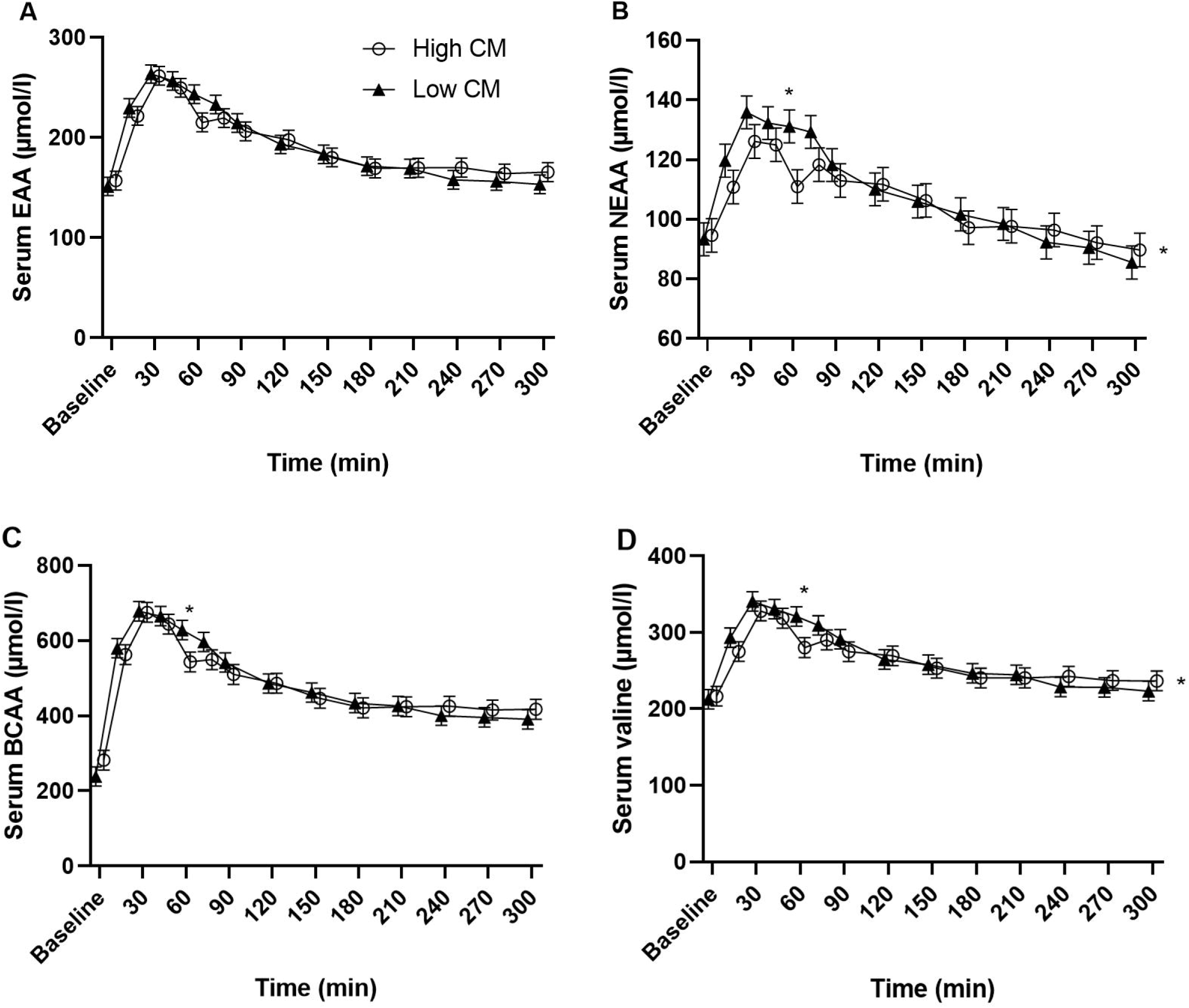
Mean ± SEM of serum essential amino acid (A), non-essential amino acid (B), branch chained amino acids (C) and valine (D) concentrations after low and high CM milk ingestion. *p < 0.05 placed above the value denotes a significant time point, at the right of the graph it denotes a significant treatment effect.

Analysis of the first 90 min, when the effect of a reduced casein coagulation is to be expected, showed higher total serum AA for low CM for both EAA (MD 45.3 µmol/L, 95% CI [14.9 – 75.6], p=0.004 and T = 60 p = 0.001) and NEAA (MD 56.7 µmol/L, 95% CI [24.7 – 88.6], p<0.001, T = 60, p = 0.005).

Relative to high CM milk, low CM milk showed earlier appearance of AAs more dominant in casein, such as proline (MD 4.18 µmol/L, 95 % CI [2.38-5.98], treatment p < 0.001), while there was no difference in AA appearance of AAs more dominant in whey protein, such as leucine (MD 0.563 µmol/L, 95 % CI [-0.99-2.12], treatment p = 0.477). In line with these observations, analysis of the estimated serum casein/whey AA ratio showed a difference between the treatments (MD 0.056, 95% CI [0.022-0.091], treatment p = 0.002, T = 30 min p = 0.015) (**Figure 5**).

**Figure 5.**
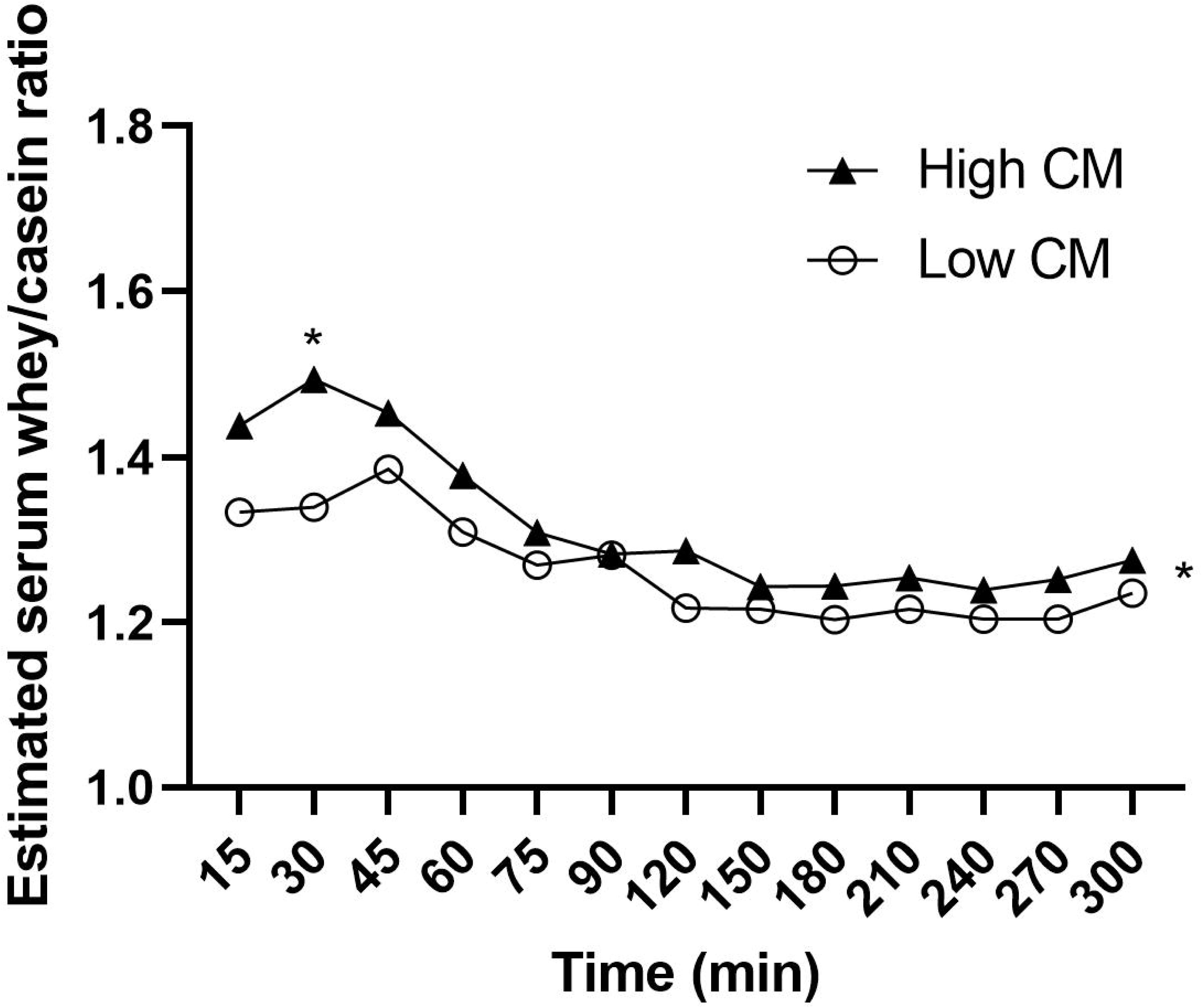
Estimated serum whey/casein AA ratio over time after low and high CM milk ingestion. *p < 0.05 placed above the value denotes a significant time point, at the right of the graph it denotes a significant treatment effect.

### Glucose and insulin

Glucose over time did not differ between treatments (MD -0.047 mmol/L, 95% CI [-0.373, 0.278], p = 0.915) and there was no time*treatment interaction (p = 0.99). The insulin response over time was significantly lower for the high CM milk (MD 0.072 (mIU/L), 95% CI [0.019, 0.125], p = 0.008). Post-hoc t-tests showed that this is driven by time points T = 30 (MD 8.4, 95% CI [2.8, 14.0], p = 0.004) and T = 45 min (MD 6.9, 95% CI [1.3, 12.5], p = 0.016). There was no time*treatment interaction (p = 0.38). Graphs of both insulin and glucose can be found in **Figure 6**.

**Figure 6.**
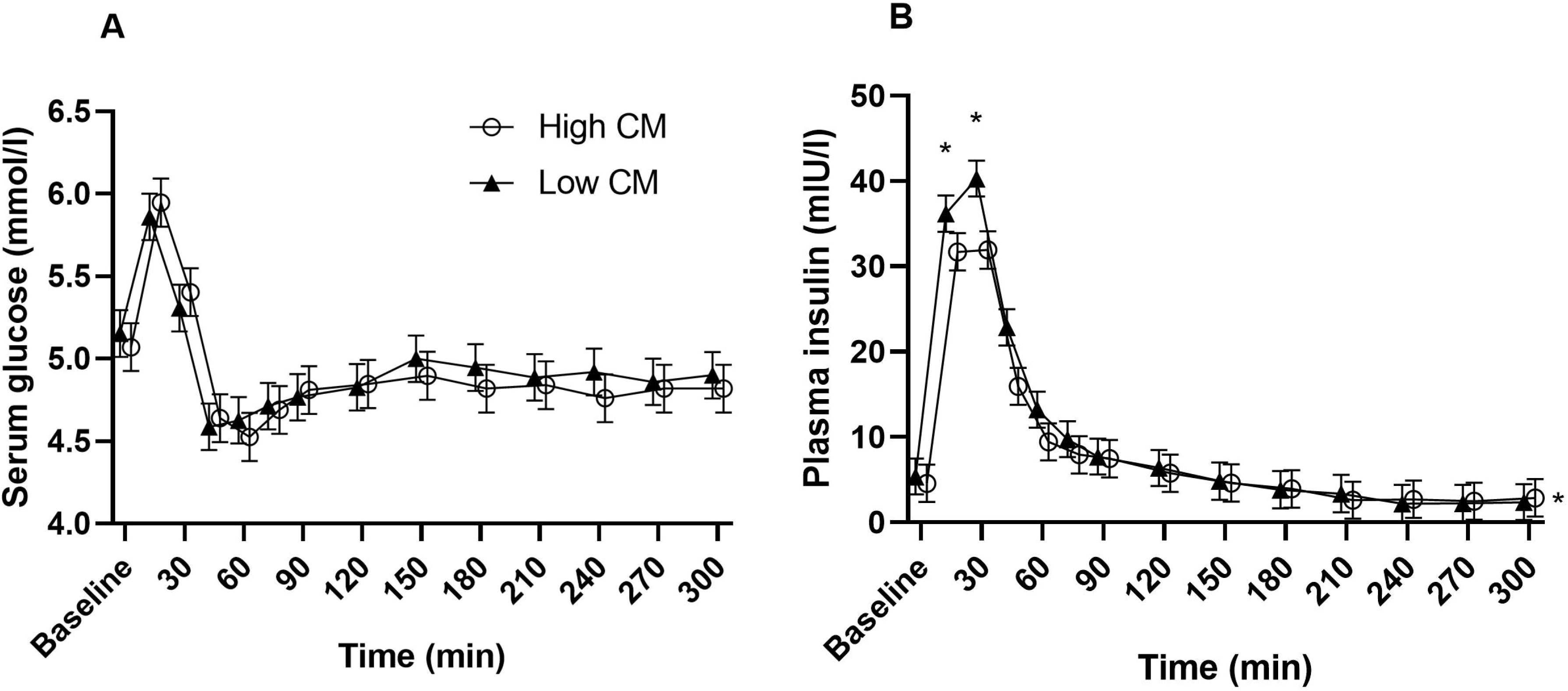
Mean ± SEM plasma glucose (mmol/L) and serum insulin (mIU/L) concentrations over time after low and high CM milk ingestion. *p < 0.05 placed above the value denotes a significant time point, at the right of the graph it denotes a significant treatment effect.

### Appetite ratings

Hunger (MD 2.7, 95% CI [-0.418, 5.87], p = 0.089), fullness (MD 0.97, 95% CI [-2.06, 3.99}, p = 0.53), desire to eat (MD 0.26, 95% CI [-2.80, 3.32], p = 0.87), prospective consumption (MD -1.22, 95% CI [-3.99, 1.56], p = 0.388), thirst (MD -2.56, 95% CI [-5.96, 0.84], p = 0.139) did not differ between treatments (**Supplementary figure 10**). There were no interaction effects for any rating.

### Time effects

There was a significant time effect for all mixed model analyses of AA, GE, glucose, and insulin, coagulation metrics and appetite ratings (all p < 0.001).

## DISCUSSION

To the best of our knowledge this is the first *in vivo* study that directly evaluated the effect of milk mineral composition on gastric casein coagulation, GE, AA and glycemic responses. Among the image texture metrics used to estimate the degree of coagulation, homogeneity and busyness were higher for the high CM milk compared to the low CM milk. As expected, the rate of gastric emptying, as measured by gastric volume curves over time and GE-t50, was similar for the two milks since they only differed in mineral composition. The serum AAs that are more dominant in casein, such as proline, and estimated serum casein/whey AA ratio were significantly higher for the low CM milk, predominantly driven by time point T = 60 min. There was no difference in glucose response. The insulin response was significantly lower for the high CM milk: driven by time points T = 30 and T = 45 min which were in line with differences in postprandial BCAA valine.

We used image texture metrics as an objective measure to quantify the degree of coagulation. A difference in homogeneity and busyness implies a difference in coagulation between the high CM milk and the low CM milk. This is in line with previous *in vitro* work applying model IF formula and simulated infant digestion^26^ and our *in vitro* digestions of the test products under simulated adult conditions (Supplementary figure 2). Both *in vitro* studies showed a substantial difference in coagula between low and high CM samples, in line with an overall reduced casein coagulation and formation of a more open structured curd for the low CM milk. However, these measures should be further validated, since such image analyses to quantify the degree of structure formation have been used in other areas^48, 49^, but are novel for characterizing gastric protein coagulation^29^. In the current study, homogeneity and busyness were both higher, which seems contradictive: a higher homogeneity may imply a lower degree of coagulation, while higher busyness may imply a higher degree of coagulation. What should be considered is that higher homogeneity could not only reflect a homogenous liquid, but also result from the presence of large coagulates. Another aspect to consider is that not only the size, but also the structure is an important characteristic of coagula. For instance, we know that some coagulates are firm and have a dense structure, and greater weight than less dense coagulates with approximately the same volume^50^. Indeed, lower CM resulted in smaller and softer curd particles *in vitro*, likely because of higher concentration of non-micellar casein, which hinder enzymatic coagulation of casein micelles^26^. MRI is sensitive to water content, so could provide information on water contained in the coagulum and therefore its density. It should be noted that MRI image texture parameters are affected by the resolution of the input images and could detect differences in image intensity patterns that are not clearly distinguishable by viewing the MRI images. This needs further validation by concomitant analysis of MRI images and coagulates that differ in size and density. Other MRI techniques are being developed that can provide molecular-level information such as measurement of the magnetization transfer ratio and relaxation rates^42, 51, 52^. These measurements require additional MRI spectra to be recorded, but could be used in follow-up research to examine more subtle differences in protein coagulation *in vivo*.

The differences in coagulation between the two milks did not affect gastric emptying rate; the GE curves were similar for both milks. However, the threshold analysis showed that most of the liquid fraction emptied sooner than the semi-solid (coagulated) fraction. It is known from *in vitro* and animal *in vivo* models that complete breakdown and subsequent GE of the coagulated casein fraction can take longer than complete emptying of the liquid fraction^4, 7, 13, 17, 26, 53^. This is likely a consequence of the increased particle size exceeding the maximum size that can pass the pyloric filter^14^ and may be physiologically important to provide a sustained release of amino acids to the neonate. Accordingly, whey proteins, which remain soluble, have a higher GE rate than caseins^18^. Studies comparing whey and casein show a difference in overall GE as assessed with MRI and ultrasonography^17, 54, 55^. However, one study with preterm infants using whey and casein-dominant formulas found similar GE^56^. Likely, overall GE rates vary between studies likely as a consequence of the different formulations used.

Gastric casein coagulation differences can likely explain the observed difference in overall postprandial AA profiles. Looking at the first 90 min, when the effect of differences in casein coagulation could be expected, both EAA and NEAA responses were significantly lower for high CM milk. This is in line with our hypothesis and is probably caused by the delay in emptying of the coagulated casein fraction. Indeed, proline, an AA more dominant in casein AA was significantly different between the treatments, whereas leucine, an AA that is more dominant in whey protein, was not, further illustrating that micellar calcium only affects the coagulation and digestion of the casein fraction. This is strengthened by the findings of differences in the estimated serum whey/casein AA ratio. A reduced casein mineralization can also explain recent *in vivo* observations where overall digestion of mineral-depleted milk protein concentrate was faster than that of a regular CM milk^57^.

No differences in glucose responses were observed, which is not surprising since carbohydrate (lactose) concentration was the same for both treatments. The lower insulin levels observed in response to high CM milk are likely due to the difference in BCAAs and/or other insulinotropic AAs present in the milk. In this study the BCAA valine was significantly higher for the low CM milk. The BCAA have the potential to influence insulin responses^58, 59^.

In conclusion, milks with different mineral compositions show different coagulating properties, as measured with higher serum AA in low CM milk, confirming *in vitro* results. Coagulation differences were further supported by MRI image analyses. Although the different coagulation properties did not influence overall GE the liquid fraction emptied quicker, while the coagulum persisted. This is in line with the difference in AA kinetics where the effects were predominantly driven by AAs more dominant in casein. The results suggest that the mineral composition of milk can influence gastric coagulation and protein digestion. This knowledge may help to determine the optimal processing of dairy products and their effect on digestion and health. Future studies should focus on improving measurements of the degree of coagulation and coagulum structure with MRI and examining the physiological relevance of the observed differences.

## Data Availability

All data produced in the present study are available upon reasonable request to the authors

## ACKNOWLEDGEMENTS

TL, and PS designed the research; EE and JR conducted the research. TL and TH provided essential materials. EE analyzed the data and drafted the paper. EE, JR, GC, TL, TH, and PS revised the manuscript critically for important intellectual content. PS had primary responsibility for the final content. All authors read and approved the final manuscript. This study was funded by FrieslandCampina and TL and TH are employed by FrieslandCampina. All other authors declare no further conflict of interest. We thank Lisa van den Berg, Caya Lindner, Jinke Oosterhof, and Jesper Rietmeijer for assisting with data collection, Jacques Vervoort and Sebas Wesseling for their work on the AA analysis and Christophe Nioche for his support with the LIFEx software. The use of the 3T MRI was made possible by WUR Shared Research Facilities.

## List of abbreviations

95% CI: 95% confidence interval
BMI: Body mass index
GE: Gastric emptying
GE-t50: Gastric emptying half time
CM: Casein mineralization
MD: Mean difference
MRI: Magnetic resonance imaging

