## Supplementary figures for "Gastric coagulation and postprandial amino acid absorption of milk is affected by mineral composition: a randomized crossover trial"

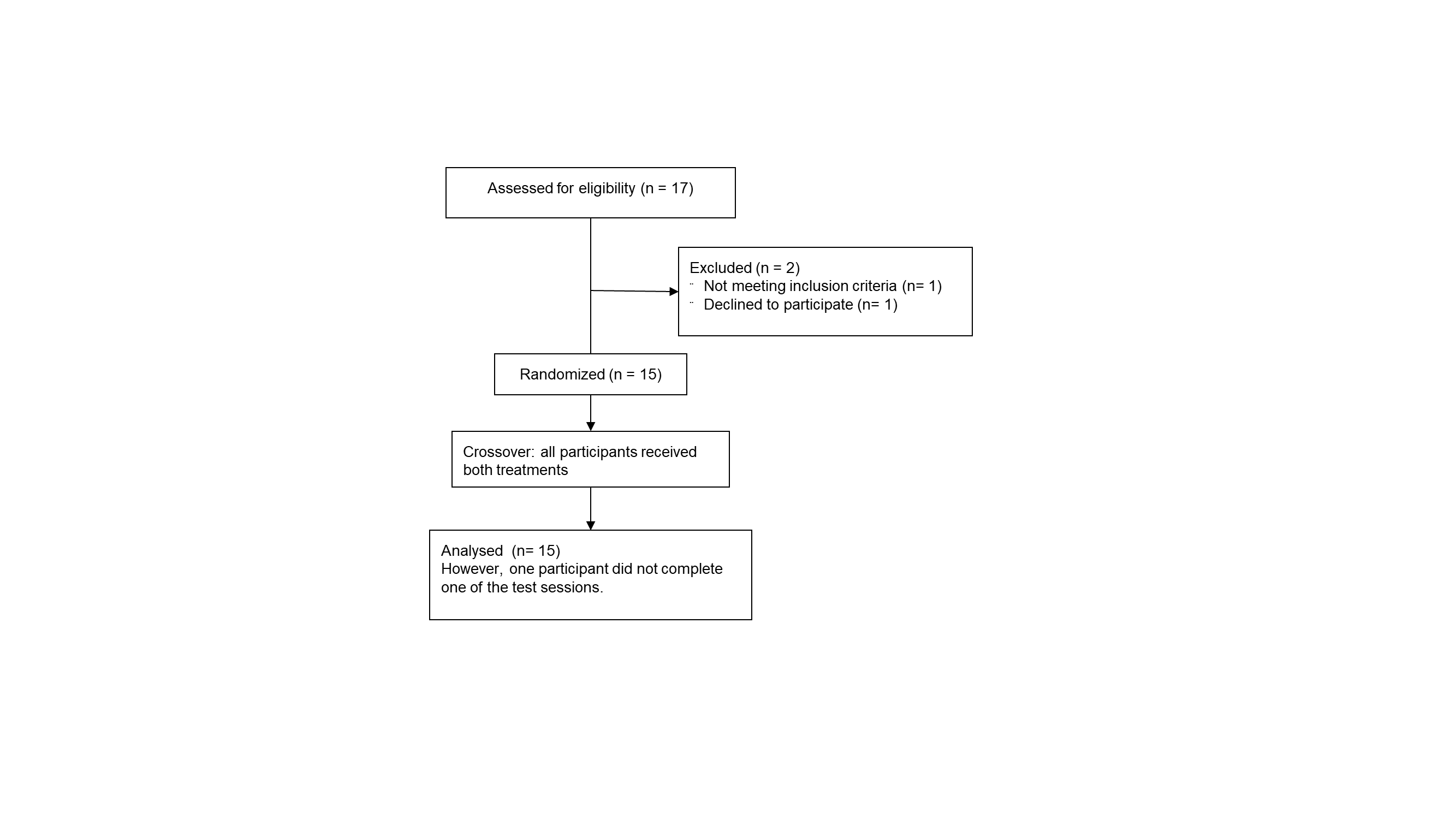

**Supplementary figure 1.** Study flow diagram

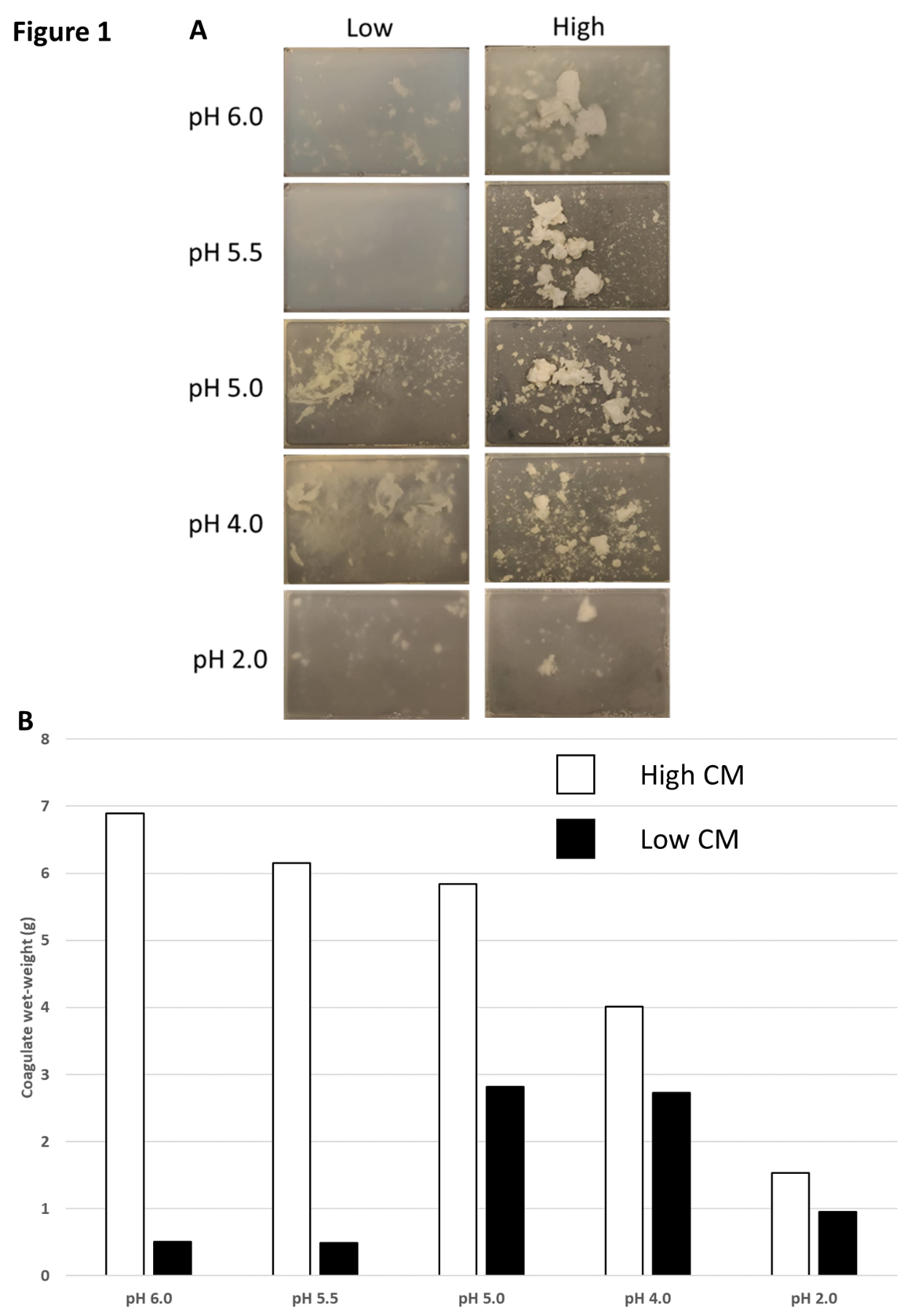

**Supplementary figure 2.** Coagulation *in vitro*

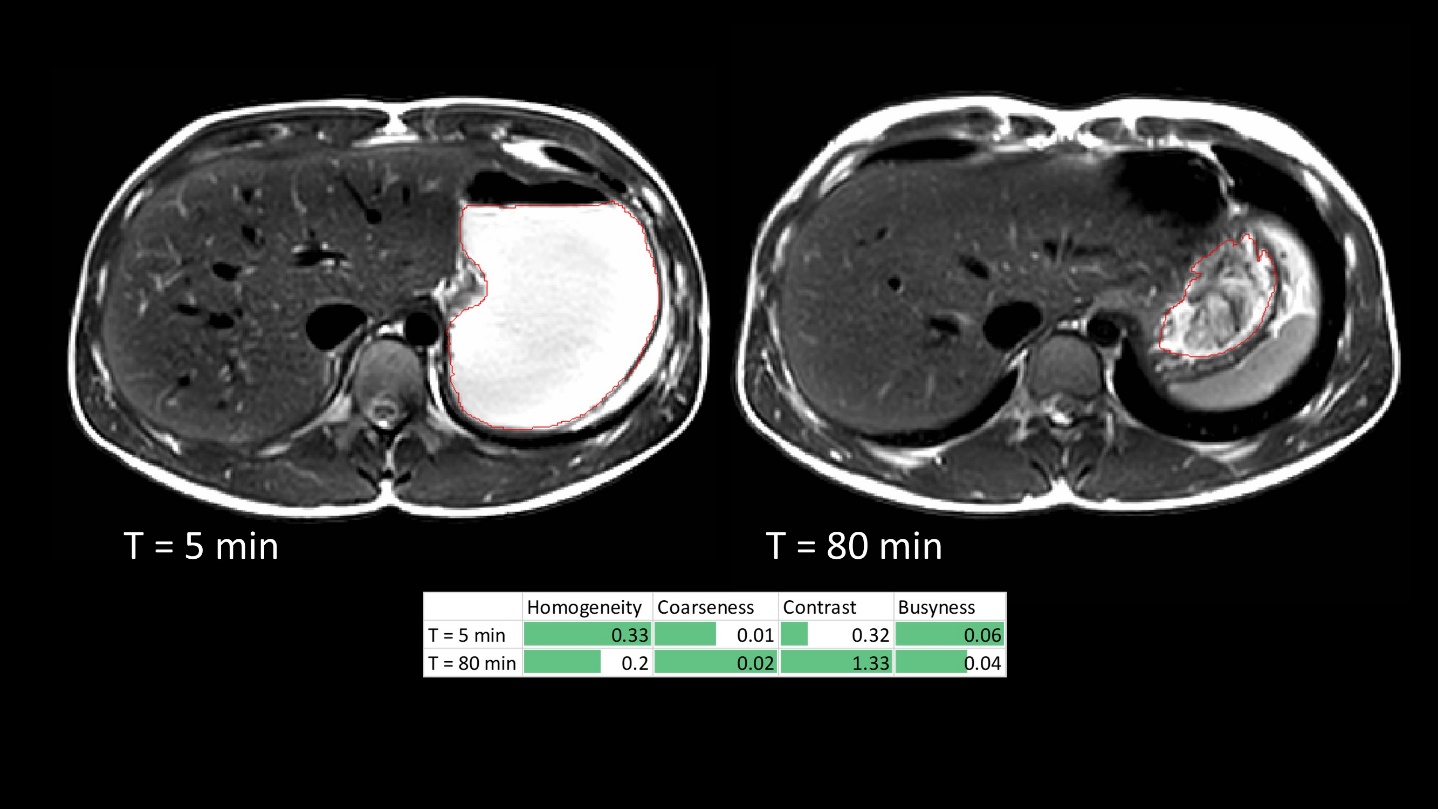

**Supplementary figure 3.** Examples of T_2_-weighted magnetic resonance images showing cross-sections of non-coagulating stomach content at T = 5 minutes and coagulating stomach content at T = 80 minutes and the associated image texture metrics.

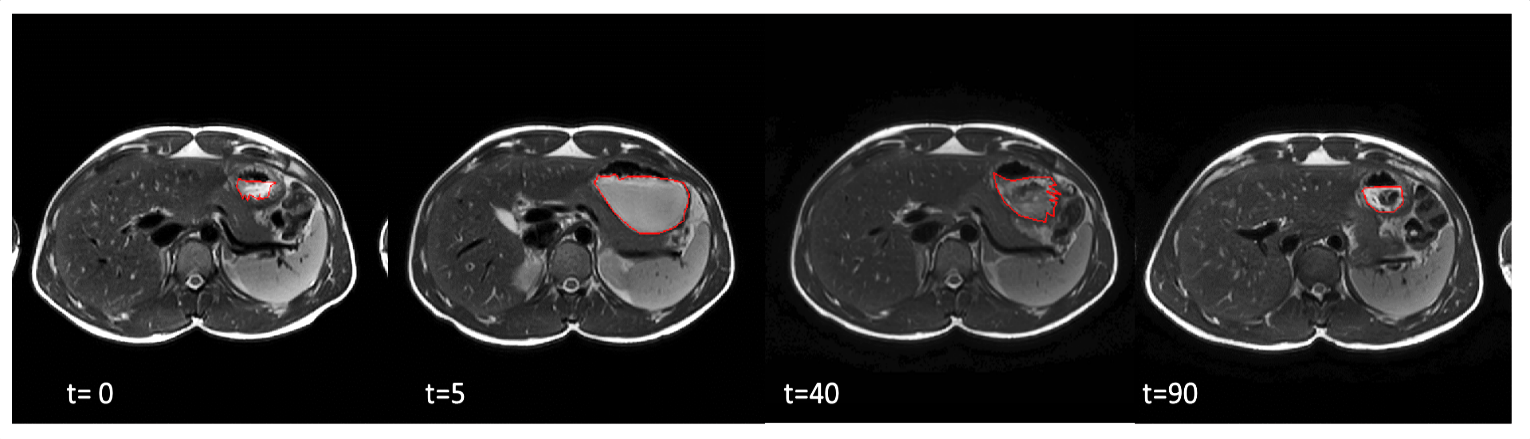

**Supplementary figure 4.** Examples of T_2_-weighted magnetic resonance images showing cross-sections through an empty stomach after an overnight fast (baseline) and after 600 ml skimmed milk consumption. The red line delineates stomach content. At T = 40 and 90 minutes milk protein coagulation can be observed by darker and lighter voxels.

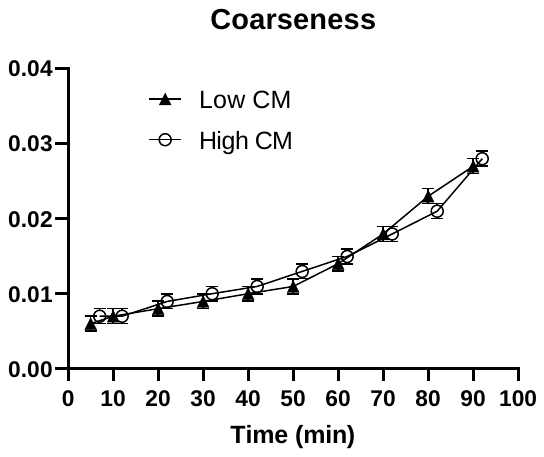

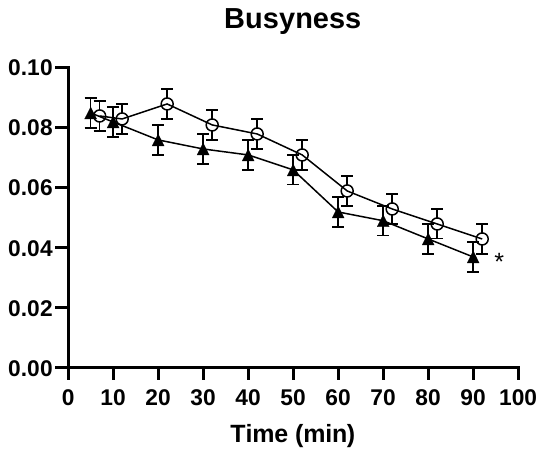

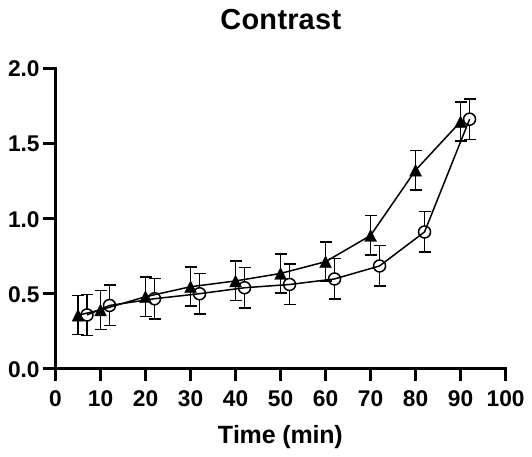

**Supplementary figure 5.** Mean ± SEM of image texture metrics coarseness, busyness and contrast. *Denotes a significant difference between treatments (n=15).

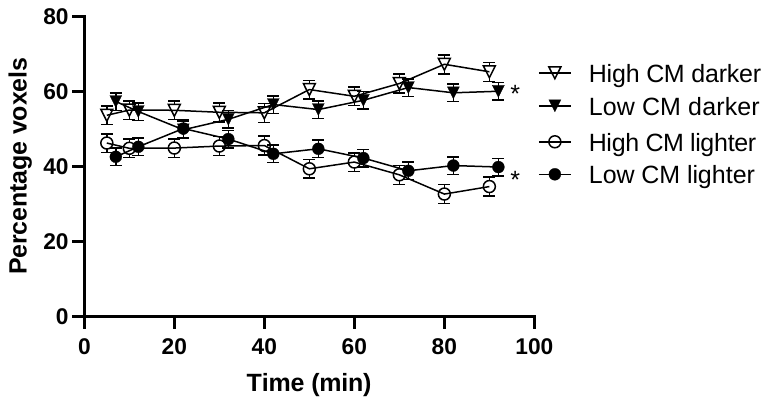

**Supplementary figure 6A.** Mean of percentage of two intensity categories of voxels of stomach content (lighter (more liquid) and darker (more solid)) after applying the thresholding method. *Denotes a significant difference between treatments (n=15).

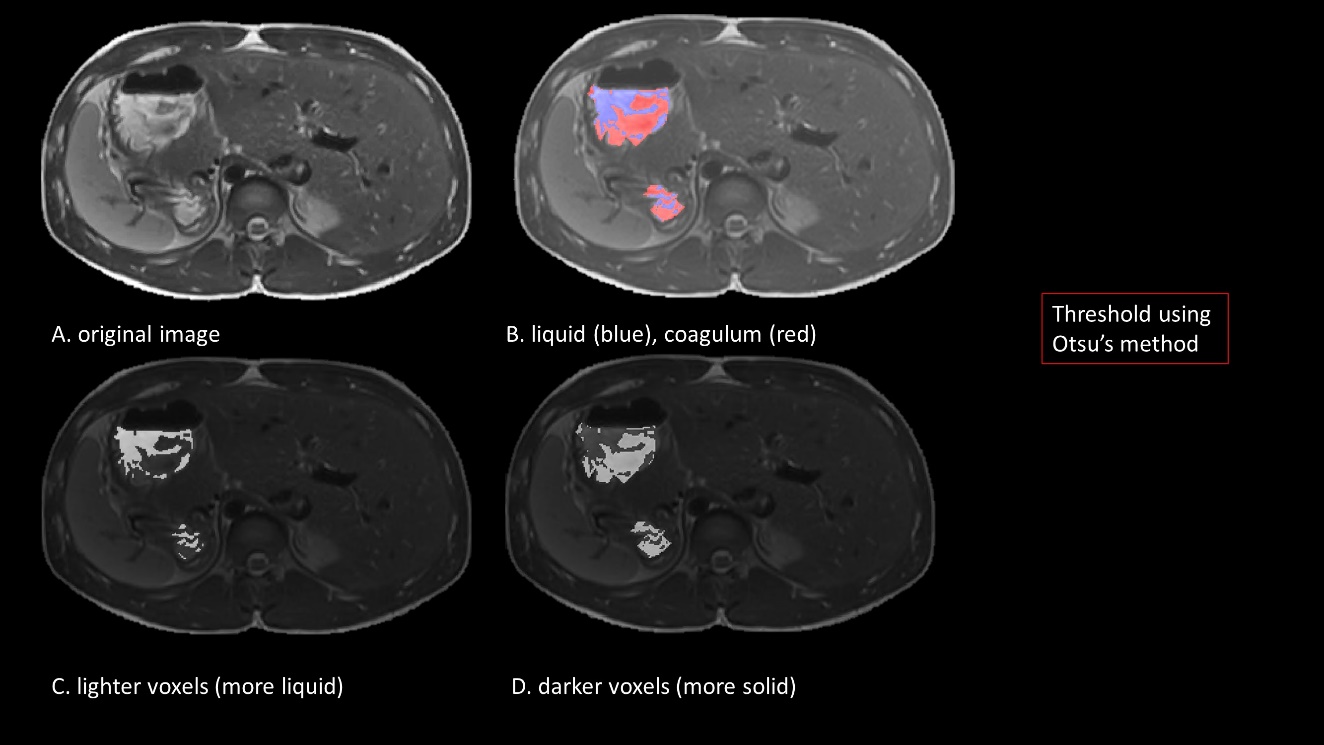

**Supplementary figure 6B.** Examples of T_2_-weighted magnetic resonance images showing cross-sections through a stomach at T = 60 min with A showing the original image and B the voxels of stomach content colored: liquid as blue and semi-solid as red after applying the thresholding method.

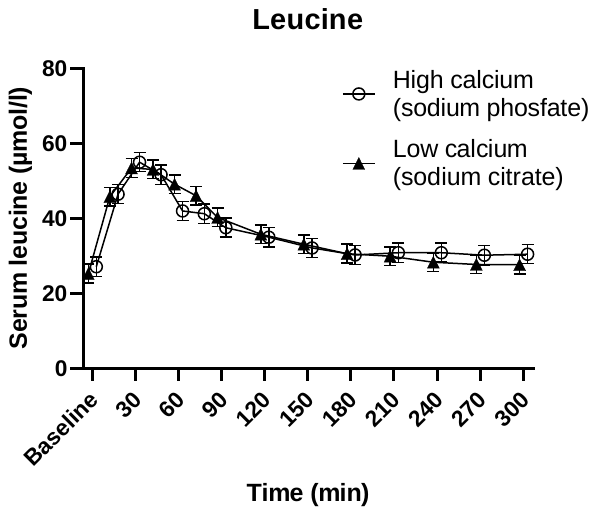

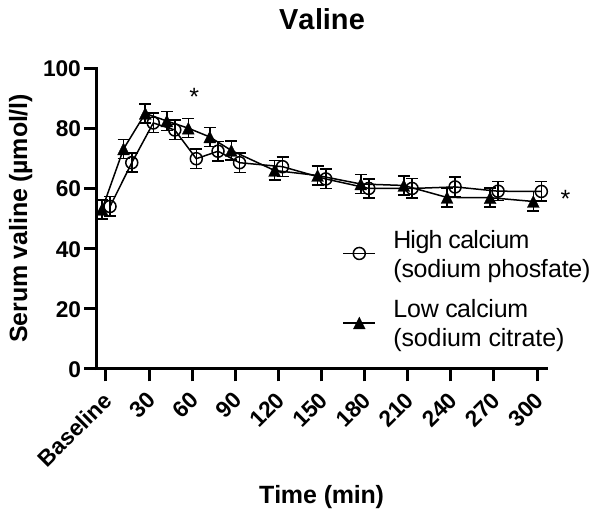

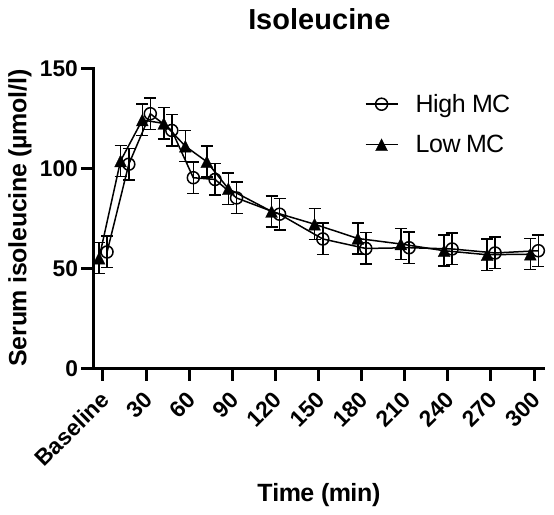

**Supplementary figure 7.** Mean ± SEM of serum branch chained amino acids over time (n=15). *p < 0.05 placed above the value denotes a significant time point, at the right of the graph it denotes a significant treatment effect.

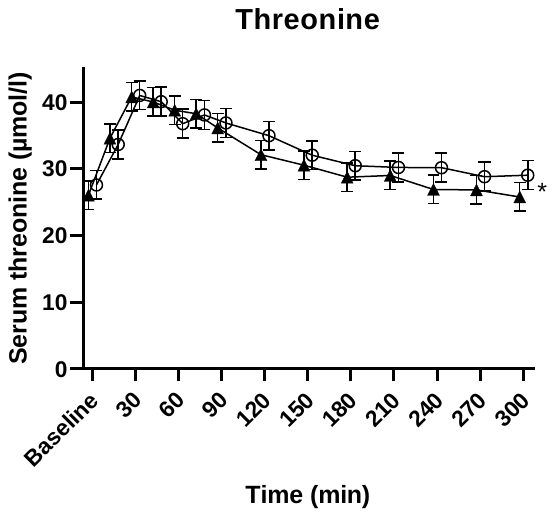

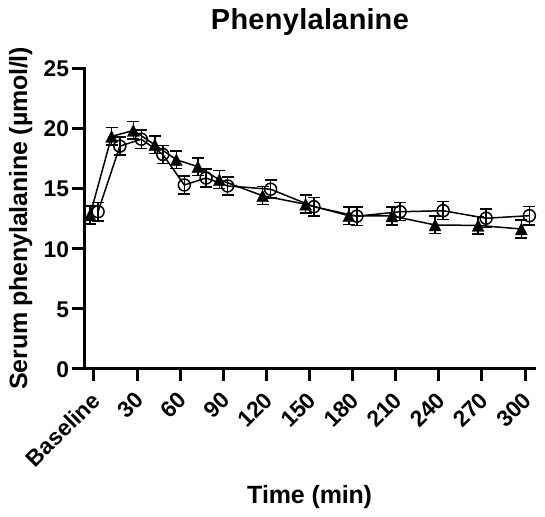

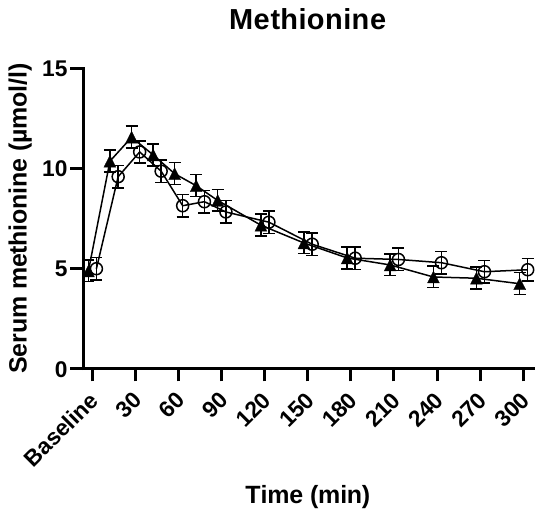

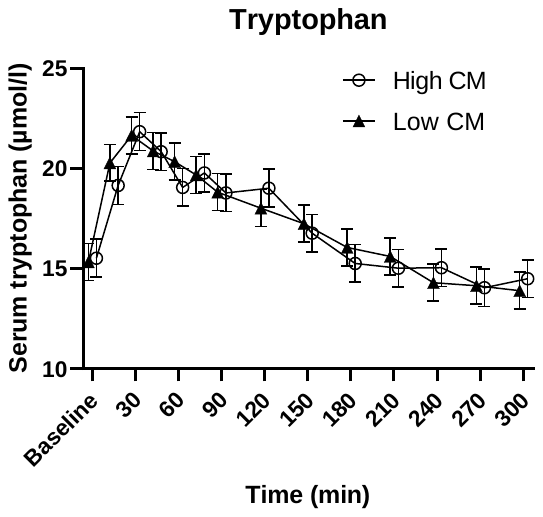

**Supplementary figure 8.** Mean ± SEM of serum essential amino acids over time (n=15). *p < 0.05 placed above the value denotes a significant time point, at the right of the graph it denotes a significant treatment effect.

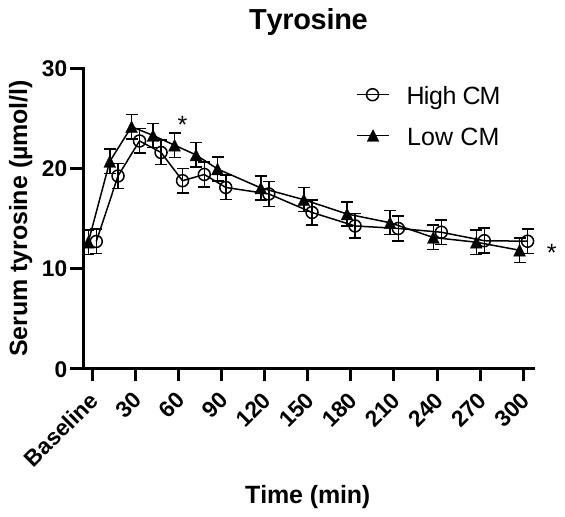

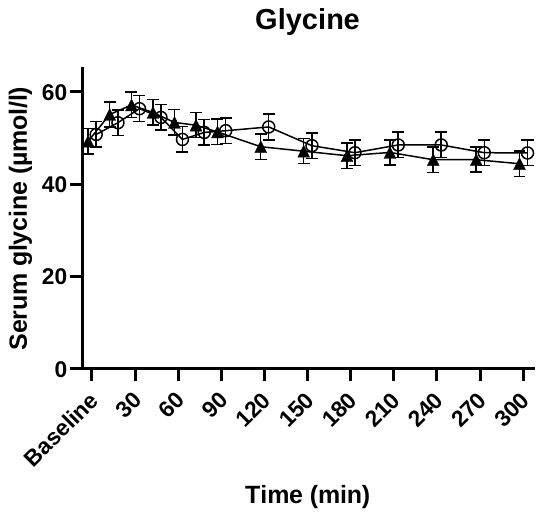

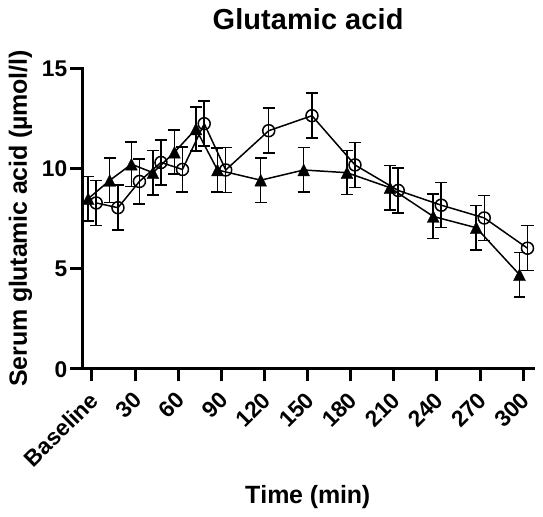

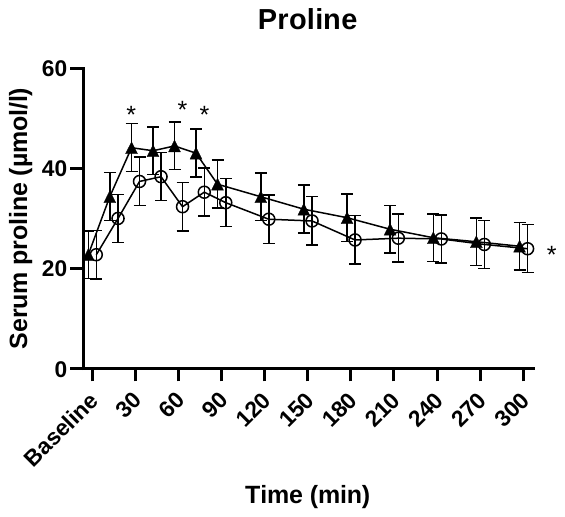

**Supplementary figure 9.** Mean ± SEM of serum non-essential or conditionally essential amino acids over time (n = 15). *p < 0.05 placed above the value denotes a significant time point, at the right of the graph it denotes a significant treatment effect.

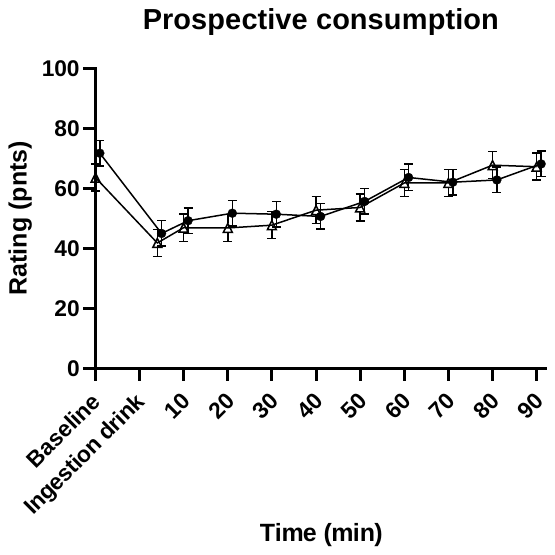

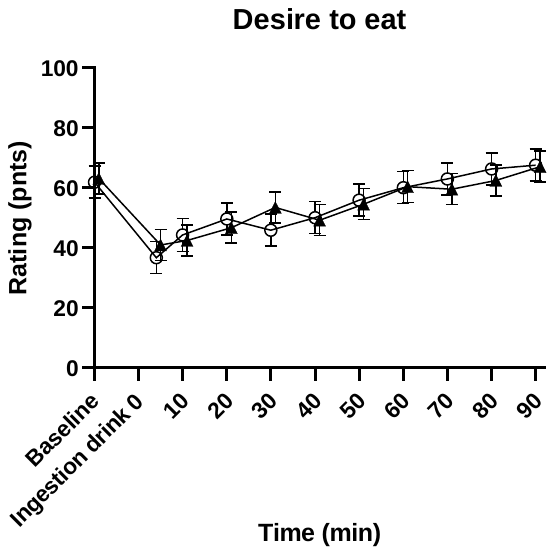

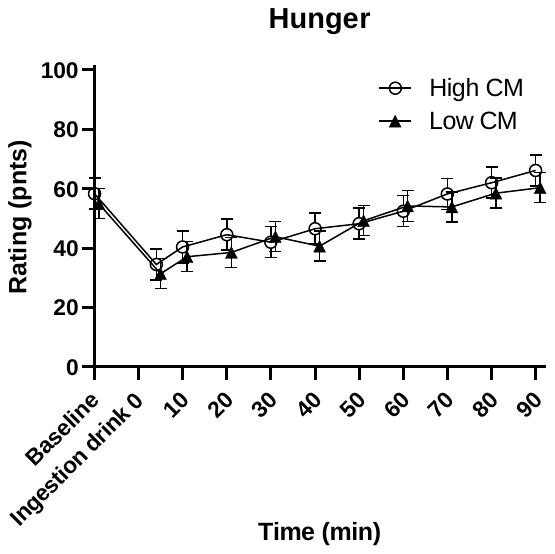

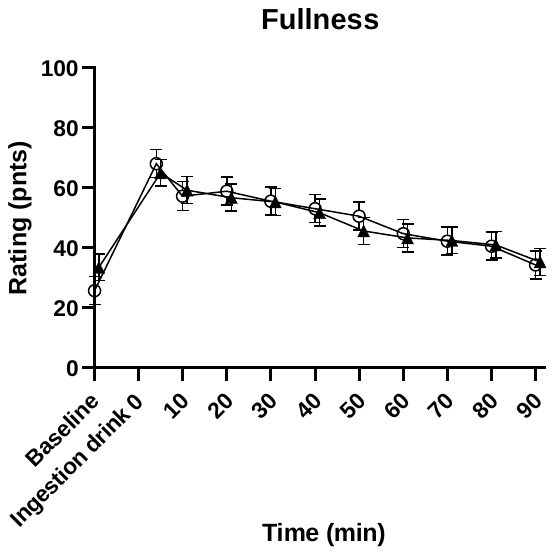

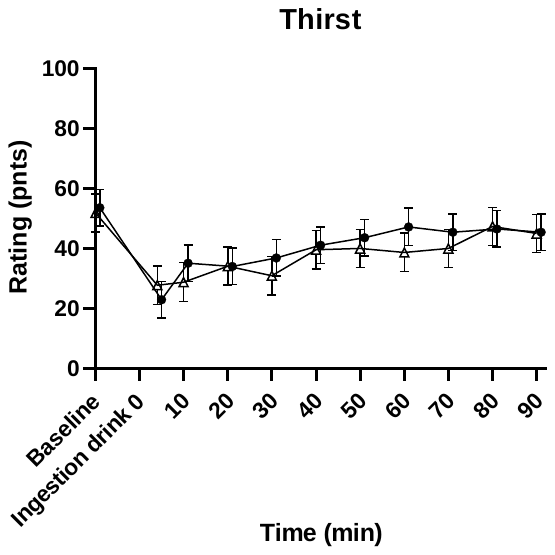

**Supplementary figure 10.** Mean ± SEM of appetite ratings hunger, fullness, prospective consumption, desire to eat and thirst (n=15).
